# Structural limits of single-barrier reform in algorithmic recourse: a formal series-system model with implications for digital health

**DOI:** 10.64898/2026.02.22.26346836

**Authors:** A.C. Demidont

**Affiliations:** Nyx Dynamics LLC, Philadelphia, Pennsylvania, United States of America

**Keywords:** algorithmic recourse, algorithmic fairness, series-system reliability model, interaction effects, global sensitivity analysis, digital health governance, algorithmic regulation, clinical AI

## Abstract

Algorithmic decision systems mediate access to healthcare, credit, employment and housing, yet individuals who experience adverse decisions face multi-stage barriers when seeking recourse. We formalize these barriers as a series-structured system with 11 empirically parameterized stages across three layers (data integration, data accuracy and institutional access) and prove that single-barrier interventions are bounded by baseline system success. Under baseline parameterization derived from federal datasets and peer-reviewed algorithmic audit studies, end-to-end recourse probability is 0.0018%. Removing any single barrier yields negligible improvement (<0.02%). Factorial decomposition reveals that the three-way cross-layer interaction accounts for 87.6% of achievable improvement, illustrated by Shapley attribution, Sobol sensitivity analysis and bootstrap resampling (n = 1,000) within prespecified perturbation ranges. Introducing dependence among the stage pass-events with a Gaussian-copula analysis (latent correlation up to ρ = 0.5), while preserving each marginal pass probability, leaves the comparative conclusion—single-layer reform yields far less than coordinated reform—intact, while a repeated-attempt topology attenuates it, delimiting the model’s domain of validity. These findings are properties of the modeled system rather than empirical measurements of real recourse pathways; they yield testable hypotheses for digital-health governance—chiefly that single-layer fairness interventions are structurally unlikely to produce large absolute gains—pending validation against observed recourse data.

**Author summary:** When an algorithm makes an adverse decision about you—denying a loan, filtering your job application, or miscategorizing your health risk—what does it take to correct it? We built a formal mathematical model of the 11 barriers a person must clear to obtain recourse, from detecting the error to accessing legal resources, and studied its structure. Calibrated with data from federal agencies and one healthcare audit—a provisional, cross-domain calibration—the model puts the probability of clearing all barriers below 2 in 100,000, and in the model no single-barrier fix raises it by more than 0.02%. This follows from the model’s multiplicative structure: barriers compound rather than add. Within the model, the three-way interaction across data integration, data accuracy and institutional access accounts for 87.6% of the achievable improvement. These are properties of the model rather than measurements of real recourse; they generate a testable hypothesis—that coordinated, multi-layer reform may outperform isolated single-layer fixes—which remains to be validated against observed recourse data.

## Introduction

Healthcare algorithms increasingly determine which patients receive timely interventions, with documented instances where a cost-for-need proxy variable systematically under-identified high-risk Black patients—correcting the proxy would have raised the share of Black patients flagged for additional care from 17.7% to 46.5% [1]. In clinical settings, algorithmic decisions include risk stratification scores, automated triage, prior authorization determinations, resource allocation and eligibility assessments; when these produce adverse outcomes, patients must navigate error detection, data correction and institutional appeals processes that we term clinical AI recourse. Analogous barrier structures arise in credit, employment and housing: approximately 75% of US employers use recruitment management systems to filter candidates before human review [2], and credit scoring algorithms shape financial access for virtually all Americans [3].

When these systems encode discrimination, individuals face multi-stage procedural frictions when seeking recourse. We treat healthcare AI as a motivating application rather than the primary empirical context: most model parameters are in fact drawn from credit, employment and housing domains, where empirical barrier data are available [4,5], and these span institutional boundaries regulated by separate agencies, creating regulatory fragmentation [6]. We therefore present the paper as a formal, cross-domain model whose healthcare-specific implications are hypotheses rather than measurements.

The sociotechnical nature of algorithmic systems means discrimination arises not from any single component but from interactions among design choices, data practices, institutional contexts and deployment environments [7]. This parallels well-characterized phenomena in reliability engineering, where series systems exhibit failure dynamics dominated by component interactions rather than individual failure rates [8,9], and in patient safety, where alignment of multiple barrier failures produces system-level adverse events [10]. Despite sustained regulatory effort targeting individual barriers— improving data accuracy, providing legal resources, mandating algorithmic audits— discrimination persists, suggesting current approaches may misunderstand the problem’s structure.

We hypothesized that barriers to algorithmic recourse function synergistically such that incremental single-layer interventions produce near-zero absolute gains unless coordinated across layers. Specifically, we ask: in a multi-stage barrier system governing recourse from adverse algorithmic decisions, when do single-barrier reforms have negligible absolute impact, and how much of achievable improvement is attributable to cross-layer interactions? We formalize this as a series-structured barrier system, prove closed-form bounds on single-barrier intervention effects and quantify the interaction structure using factorial decomposition, Shapley attribution and global sensitivity analysis. This framework provides a quantitative basis for coordinated intervention design in clinical AI governance and beyond.

## Results

### Baseline system characteristics

The 11-barrier model (Table 1) yielded a baseline success probability of 0.0018%, indicating that fewer than 2 in 100,000 individuals successfully navigate all barriers to resolve adverse algorithmic decisions.

**Table 1.**
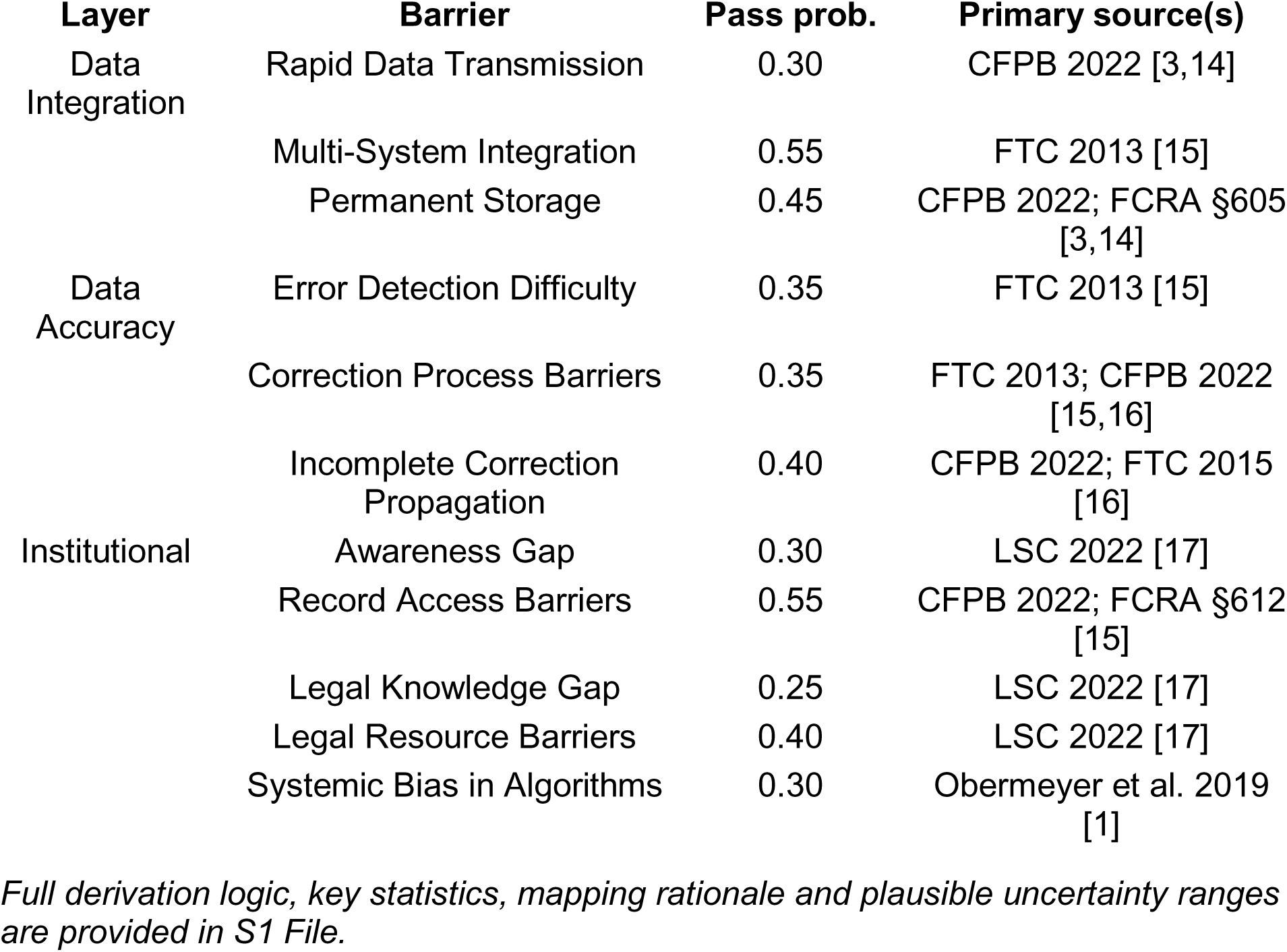
Barrier parameters, layer assignments and primary sources.

### Analytical bounds on single-barrier interventions

Proposition 1 establishes that removing barrier j yields absolute improvement Δ_j_ = P · (1/p_j_ − 1), which is linear in baseline success P. Counterfactual analysis matched this prediction: all individual barrier removal effects were <0.02%, with the maximum observed for Legal Knowledge Gap (Δ = 0.0054%). This value matches the analytical bound P · (1/0.25 − 1) ≈ 0.0054% for the barrier with lowest pass probability (Fig 1).

**Fig 1.**
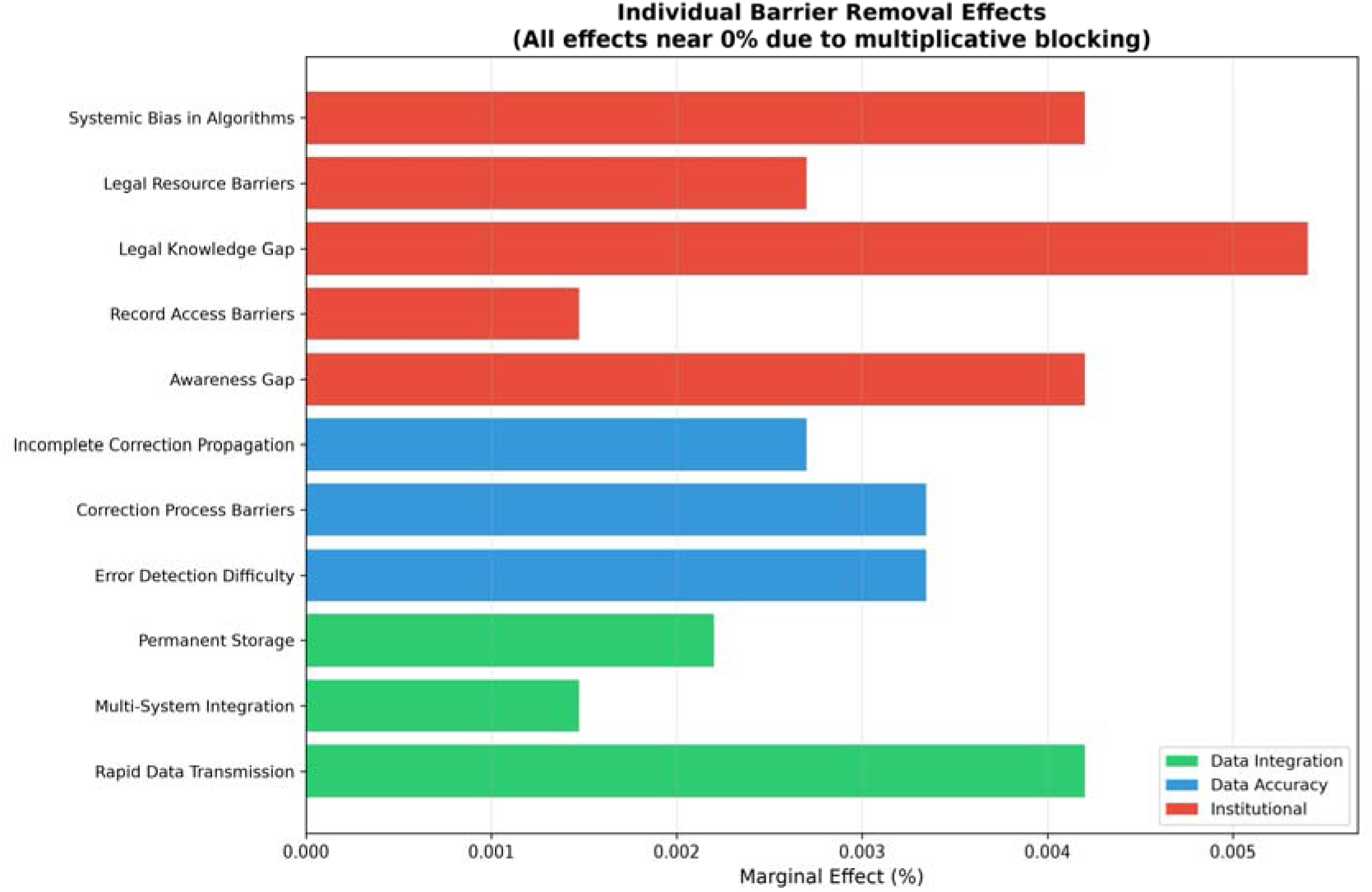
Single-barrier removal effects are structurally small. Marginal effect of removing each barrier while all others remain in place; all effects are <0.02%, with the maximum for Legal Knowledge Gap removal (0.0054%), matching the Proposition 1 bound. Error bars are 95% bootstrap confidence intervals (seeded). Reported descriptively.

### Strategy comparison

Comparison of barrier removal strategies—forward, backward, greedy by marginal impact and random ordering—revealed characteristic threshold trajectories with near-zero improvement until removal of final barriers (Fig 2). All strategies converge only at near-complete removal; removal ordering is irrelevant to this threshold behavior. We report this descriptively; the previously reported strategy-equivalence ANOVA (F = 0.07, P = 0.98) has been removed, as a comparison of three deterministic trajectories against a single averaged random trajectory lacks a defensible sampling model.

**Fig 2.**
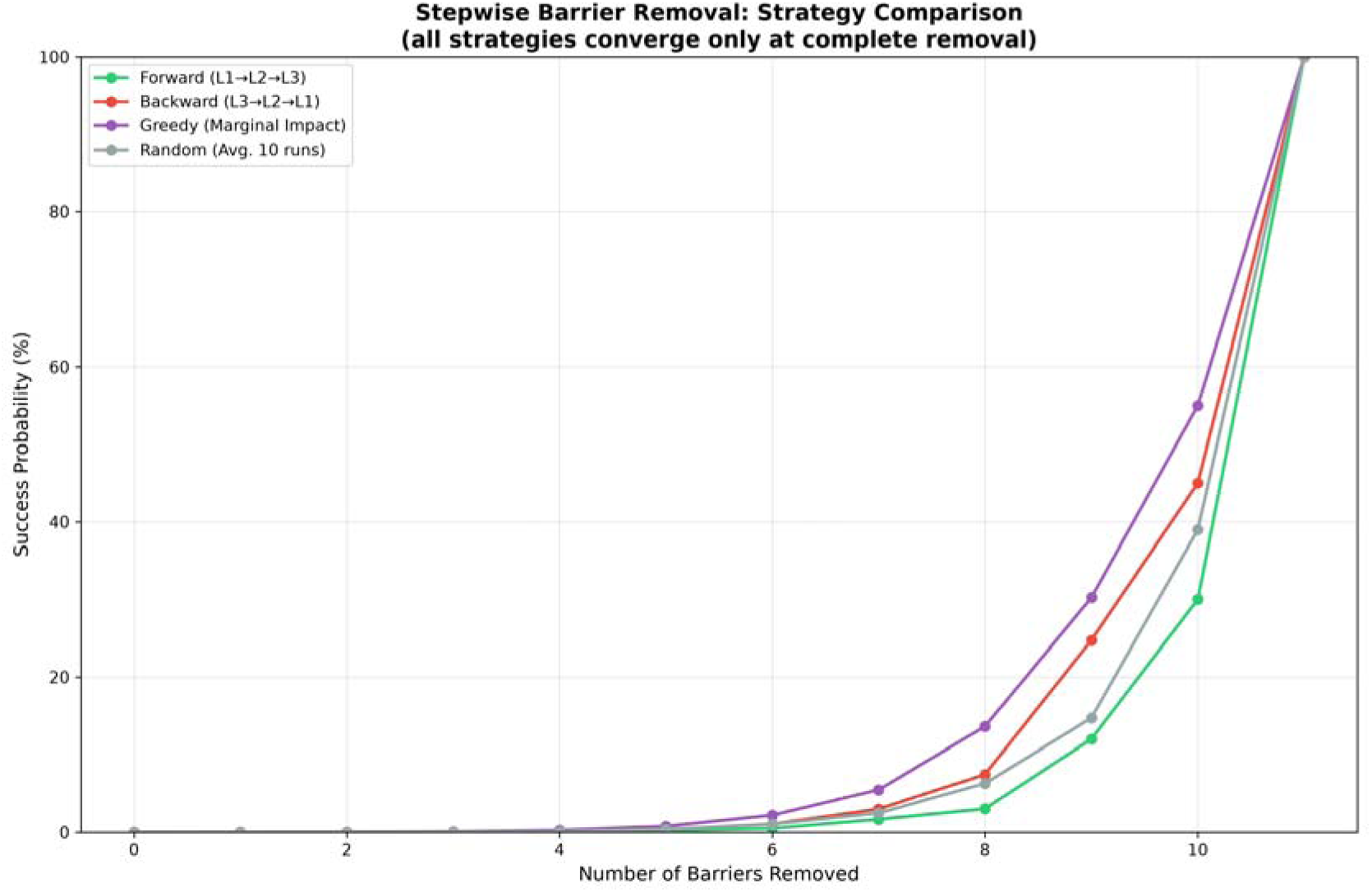
Only near-complete barrier removal helps. Forward, backward, greedy-by-marginal-impact, and random removal strategies all show threshold trajectories, with negligible improvement until the final barriers are removed. Reported descriptively; no inferential test is applied.

### Cross-layer interaction structure

Factorial analysis revealed higher-order interaction as anticipated by Proposition 2 (Fig 3, Table 2). The three-way interaction accounted for 87.6% of achievable improvement—a baseline-relative factorial share, distinct from a variance decomposition and from the smaller Sobol total-minus-first-order gaps reported below. Main effects contributed minimally: Layer 1 (0.02%), Layer 2 (0.03%), Layer 3 (0.36%). This 87.6% share is a model-derived numerical result under the calibrated parameters, not a universal property.

**Fig 3.**
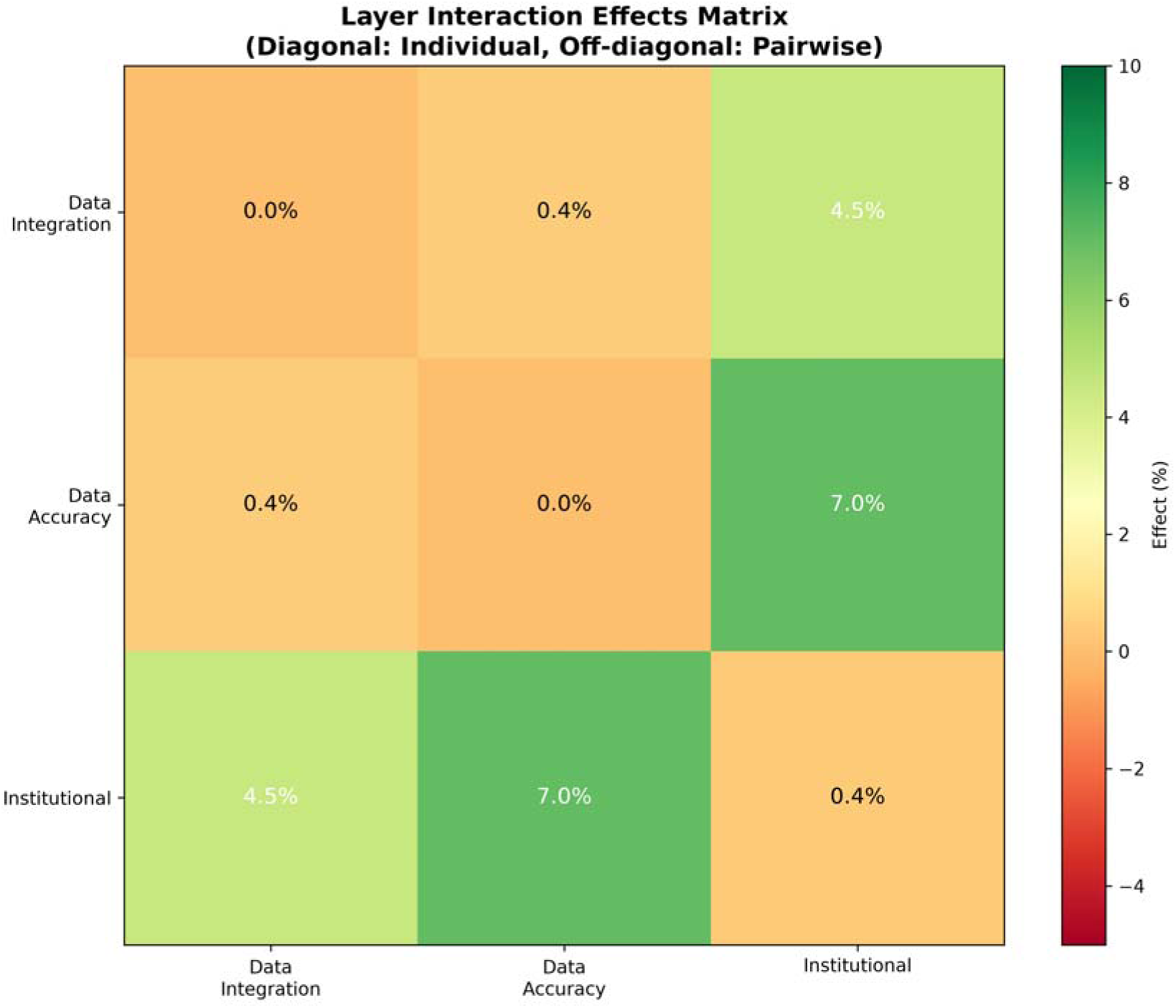
Improvement is cross-layer. Heatmap of main effects (diagonal) and pairwise interactions (off-diagonal) among the three layers. The largest pairwise term is Data Accuracy × Institutional (7.03%); the three-way interaction (87.6% of achievable improvement, not shown) dominates.

**Table 2.** Baseline-relative factorial decomposition of achievable improvement.

| Effect | Share of achievable improvement (%) |
| --- | --- |
| Layer 1 (Data Integration) | 0.02 |
| Layer 2 (Data Accuracy) | 0.03 |
| Layer 3 (Institutional) | 0.36 |
| L1 × L2 | 0.44 |
| L1 × L3 | 4.51 |
| L2 × L3 | 7.03 |
| **L1 × L2 × L3 (Three-way)** | **87.60** |
| **Total** | **100.0** |

### Sensitivity analysis and robustness

Sobol total-order indices exceeded first-order indices for all barriers, but the gaps are small and, for several barriers, within Monte-Carlo error at the current Saltelli sample size (Fig 4); the earlier characterization that total-order indices “substantially exceeded” first-order indices for all barriers is therefore not supported. Morris screening identified Legal Knowledge Gap and Systemic Bias in Algorithms as having highest nonlinear/interaction involvement. The principal findings were stable within the prespecified ±10–33% perturbation ranges (parameter-perturbation resampling, n = 1,000); robustness is not claimed under arbitrary uncertainty—the coefficient of variation of the output rises steeply with noise, reaching ∼110% at 25% and ∼129% at 30% multiplicative noise, and the signal-to-noise ratio falls below 0 dB by 25% noise (Fig 5). All stochastic analyses are seeded (seed 42) and reproducible.

**Fig 4.**
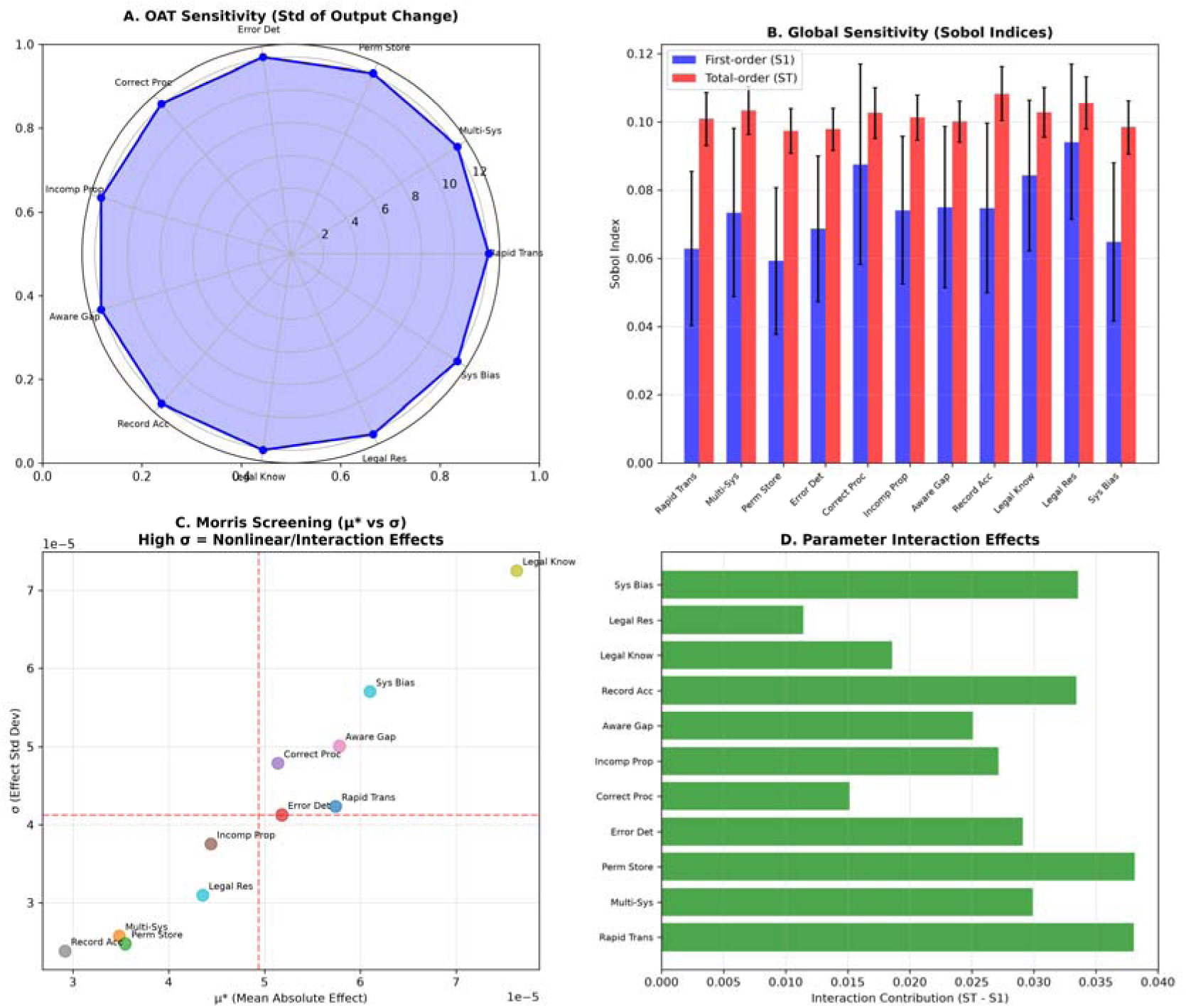
Global sensitivity. (A) One-at-a-time sensitivity radar; (B) Sobol first-order (S) and total-order (S_T) indices with 95% confidence intervals; (C) Morris screening (μ* vs σ); (D) per-barrier interaction contribution (S_T − S). Total-order indices exceed first-order indices for all barriers, modestly for several. Seeded.

**Fig 5.**
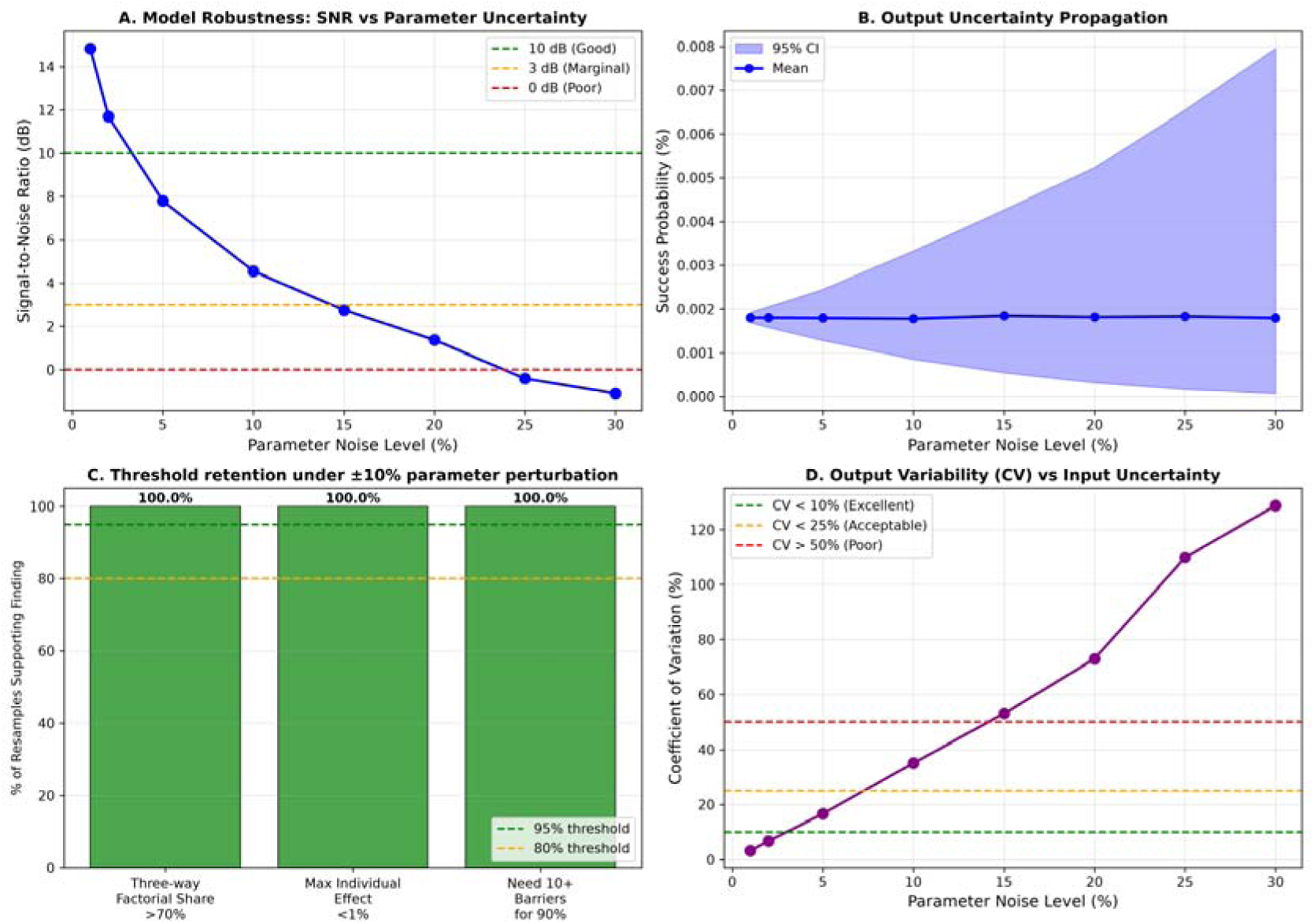
Robustness is bounded. (A) Signal-to-noise ratio vs parameter noise; (B) output-uncertainty propagation; (C) threshold retention under ±10% parameter perturbation; (D) coefficient of variation vs input uncertainty, rising to ∼110% at 25% and ∼129% at 30% noise. Stability is claimed only within the prespecified perturbation ranges. Seeded.

### Shapley value attribution

Shapley decomposition assigned fair responsibility accounting for all possible removal orderings. Top contributors (seeded, seed 42): Legal Knowledge Gap (11.0%), Rapid Data Transmission (10.7%), Systemic Bias in Algorithms (10.5%), Incomplete Correction Propagation (10.1%) and Awareness Gap (9.7%); contributions are comparatively even (no barrier >11%) and span all three layers. Unlike marginal effects—which approach zero under multiplicative blocking (Proposition 1)—Shapley values report each barrier’s average marginal contribution across all removal orderings; we treat them as a model-attributed allocation under the specified value function, not as a causal claim.

### Robustness to alternative topologies

Because the interaction structure partly reflects the assumed serial, single-pass topology, we tested two plausible relaxations (seed 42, n = 100,000; see Methods). Under a Gaussian copula coupling the 11 pass-events, positive correlation raised the baseline success probability toward the weakest barrier (from 0.0018% at ρ = 0 to 3.2% at ρ = 0.5), so the specific baseline value is correlation-sensitive; the three-way interaction share, however, remained high (77–88% across ρ [0, 0.5]), and single-lever reform remained far weaker than coordinated reform (at ρ = 0.5, the best single-layer intervention yielded 4.5% versus 96.8% for coordinated removal). Under a repeated-attempt topology (m attempts per stage), the interaction share fell to 53% (m = 2) and 25% (m = 3) as the baseline rose, indicating that the strong quantitative conclusions hold for approximately single-pass serial recourse and weaken if recourse is substantially iterative. We interpret this as delimiting the model’s domain of validity rather than invalidating it (Fig 6).

**Fig 6.**
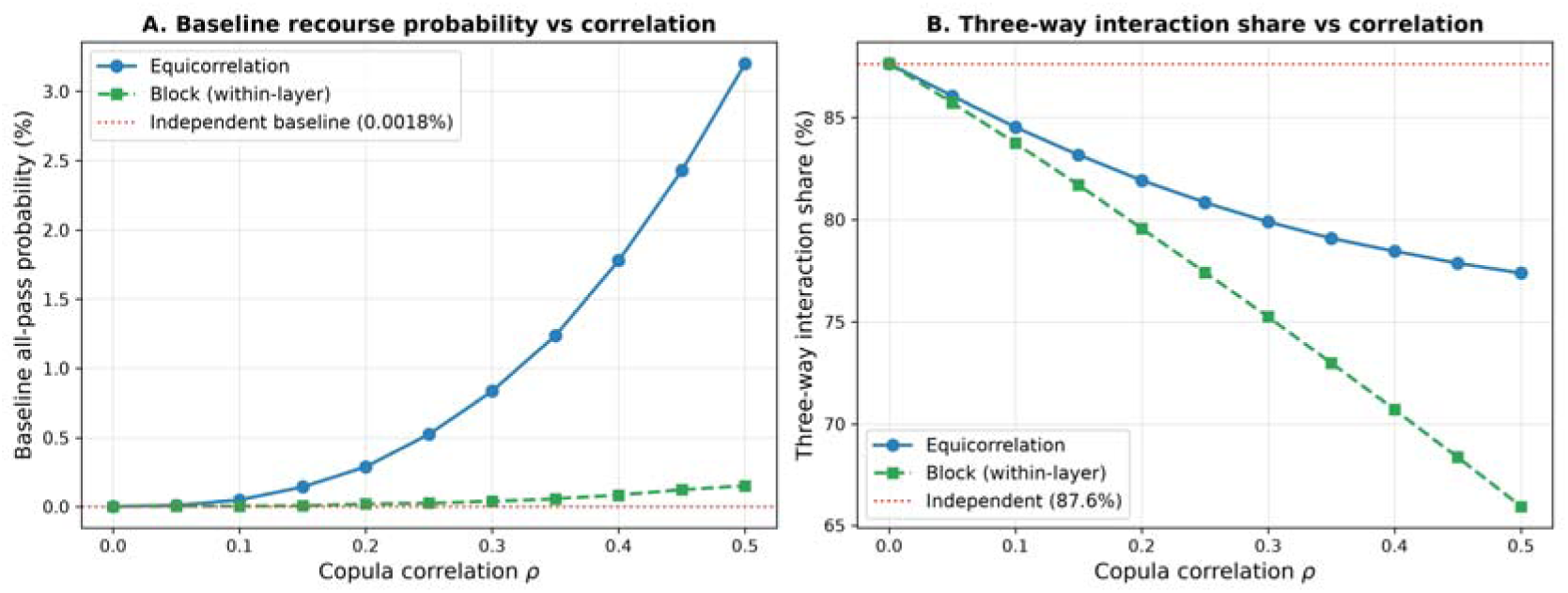
Alternative topologies delimit the model’s validity. Baseline success probability P and three-way interaction share as functions of the latent Gaussian-copula correlation ρ (equicorrelation and within-layer block variants). The comparative conclusion (single-layer coordinated reform) is robust to correlation while the baseline value is correlation-sensitive; the repeated-attempt results (m = 2, 3), which attenuate the interaction share, are reported in Supplementary Table S7. Seeded (n = 100,000).

## Discussion

Under a series-structured barrier model with cross-domain parameters, algorithmic recourse behaves as a synergistic system in which barriers reinforce each other, rendering incremental reform structurally insufficient within the model. The three-way interaction share (87.6%) offers a candidate explanation for why incremental policy reforms—including targeted improvements to data accuracy, legal access and algorithmic auditing—may have limited impact: if recourse behaves as a low-baseline series system, targeting any subset of barriers leaves the multiplicative blocking structure substantially intact. We frame this as a hypothesis rather than a demonstrated policy fact, because the model does not observe real recourse programs.

The closed-form bound established in Proposition 1 makes precise what has been intuitive in advocacy contexts: when system-level success probability is extremely low, no single-barrier intervention can produce meaningful absolute improvement. This is an algebraic property of series systems, not contingent on specific parameter values. When health systems propose that improving algorithmic audit quality or adding patient notification requirements will substantially reduce algorithmic harm, Proposition 1 demonstrates that these claims are mathematically bounded by baseline success probability regardless of the magnitude of the barrier addressed.

The distinction between probability-scale and log-scale analysis (Proposition 2) is substantively important for clinical AI governance. On the log scale, barriers contribute additively—a framing suggesting incremental reform could accumulate benefits. On the probability scale, where patient outcomes and equity metrics are evaluated, multiplicative stacking produces threshold behavior: improvement remains negligible until most barriers are removed, then increases rapidly. This threshold pattern is a generic property of series systems [8] and predicts that single-lever fairness interventions will yield small absolute effects whenever baseline recourse probability is low.

The synergistic structure parallels combination therapy in infectious disease, where incomplete antiretroviral therapy is insufficient because the virus exploits any untargeted mechanism [11]. Similarly, incomplete policy intervention addresses visible symptoms while leaving the underlying multiplicative structure intact. For healthcare AI specifically, this implies—as a hypothesis pending empirical validation—that fairness interventions limited to the algorithmic layer (bias mitigation, audit requirements) would be expected to have limited population-level impact without simultaneous attention to data integration practices and institutional access to recourse mechanisms [22,23]. The regulatory fragmentation governing algorithmic systems—CFPB for credit reporting, EEOC for employment, HHS for healthcare algorithms—mirrors the barrier fragmentation in our model and may structurally impede the coordinated approach our analysis indicates is necessary.

Interaction-dominant barrier structures likely extend to other clinical AI domains where administrative, technical and institutional processes create series-structured requirements for patient recourse. Sociotechnical systems in which fairness depends on alignment of multiple organizational and technical components [7] may exhibit analogous dynamics. Recent work on clinical AI fairness frameworks [12] and bias recognition strategies [13] has emphasized multi-level approaches; our quantitative framework provides formal support for this intuition and a measurement methodology for evaluating whether proposed interventions address sufficient layers of the barrier system.

The empirical foundation most often cited for algorithmic bias in healthcare, Obermeyer et al. [1], is a single audit of one widely used commercial risk-prediction algorithm applied to one health system. The bias stemmed from a specific design choice— predicting health-care cost as a proxy for health need—so that Black patients at a given risk score were sicker and under-referred; correcting the proxy would raise the share of Black patients flagged for additional help from 17.7% to 46.5%. This finding is compelling but context-specific and cannot, on its own, calibrate healthcare algorithmic bias at population scale; we accordingly treat it as one motivating example rather than an empirical anchor for the Systemic Bias barrier. Healthcare-AI fairness research has since matured beyond single-algorithm cost-proxy audits: chest-radiograph classifiers systematically underdiagnose under-served subgroups [24], models can recover self-reported race from medical images through latent pathways clinicians cannot detect [25], and dataset-standard and reporting frameworks have emerged to address such harms [26,12,13]. Algorithmic bias in healthcare is thus multi-mechanism and modality-spanning, and our single healthcare-derived parameter should be read in that light.

Several limitations should be noted. We note that the simulation results are consistent with but not independent of the analytical propositions, as both derive from the same multiplicative model structure. This model is a formal abstraction and does not directly observe individual-level resolution trajectories. Barrier probabilities are derived or mapped (when direct measurement is unavailable) from heterogeneous empirical sources rather than a single integrated dataset; however, each parameter is traceable to publicly available federal reports or peer-reviewed studies (Table 1 in the main text and the Supporting Information), with explicit derivation logic documented in the repository. The principal conclusions are analytical properties of the series structure (Propositions 1–2) that hold across wide parameter perturbations, consistent with parameter-perturbation resampling and global sensitivity analysis. The sensitivity analysis indicates only bounded stability within the prespecified perturbation ranges; we do not claim robustness under arbitrary uncertainty, and true uncertainty may exceed these bounds. Writing recourse as a product does not itself assume statistical independence—by the chain rule the pass probabilities are properly conditional; the genuine limitation is that the implemented parameters are largely marginal or proxy statistics standing in for conditional stage-transition probabilities. We tested the consequences directly (Robustness to alternative topologies): moderate cross-barrier correlation preserved the comparative conclusion but made the baseline value correlation-sensitive, and a repeated-attempt topology attenuated interaction dominance, delimiting the model’s domain of validity to approximately single-pass serial recourse. The model does not capture feedback loops or heterogeneity across populations. Future work should integrate longitudinal administrative data—including healthcare claims, appeal outcomes and algorithmic audit records—to empirically estimate joint barrier transition probabilities and validate the series-system structure against observed recourse outcomes.

In summary, under a series-structured barrier model, algorithmic recourse exhibits synergistic dynamics that render incremental reform insufficient in isolation within the model. The three-way interaction provides a mathematical basis for the hypothesis that coordinated, multi-layer intervention outperforms isolated reform. Under low-baseline, approximately serial recourse, the model predicts that effective mitigation may require simultaneous intervention across data integration, data accuracy and institutional access barriers rather than isolated single-layer reforms—a prediction to be tested against observed recourse data.

## Materials and methods

### Barrier model specification

We modeled algorithmic discrimination as requiring successful passage through 11 sequential barriers organized into three layers (Table 1). Layer 1 (Data Integration) comprises barriers related to speed and breadth of adverse data propagation across interconnected systems. Layer 2 (Data Accuracy) comprises barriers to detecting, correcting and propagating corrections of erroneous data. Layer 3 (Institutional) comprises barriers to awareness, record access, legal knowledge, legal resources and algorithmic bias in decision systems. We define successful recourse as full end-to-end resolution—the underlying data are corrected, the adverse decision is reversed, and the correction propagates across the relevant institutions; this is a stringent endpoint and is itself a modeling choice, since less stringent endpoints (e.g., correction at a single institution) would raise the baseline. We also note that “Systemic Bias in Algorithms” is retained in the institutional layer as the persistence of bias during recourse; a sensitivity analysis removing/reclassifying it as upstream (a 10-barrier variant) leaves the qualitative conclusions unchanged (baseline 0.0060%, three-way share 86.6%, maximum single-barrier effect <0.02%).

#### Scope of claims

We distinguish four levels of claim and apply the distinction throughout: (1) theorem-level results (Propositions 1–2), certain given the model; (2) numerical/model illustrations—the 0.0018% baseline, the 0.0054% maximum single-barrier gain, the 87.6% three-way share, and the Shapley/Sobol/Morris/bootstrap outputs—which are consistent with, but not independent confirmation of, the propositions because both derive from the same multiplicative structure; (3) empirical calibration assumptions (the 11 pass probabilities, provisional and transport-limited); and (4) real-system claims about actual recourse behavior or policy, treated as hypotheses pending validation against observed recourse data.

### Empirical parameter derivation

Pass probabilities were derived or mapped (when direct measurement is unavailable) from publicly available federal datasets and peer-reviewed studies, with explicit uncertainty ranges. Mapping decisions were conservative, favoring higher pass probabilities where ambiguity existed to avoid overstating interaction effects. Layer 1 probabilities were derived from CFPB and FTC data on credit reporting system dynamics [3,14]. Layer 2 probabilities were derived from FTC and CFPB dispute outcome data [15,16]. Layer 3 probabilities were derived from Legal Services Corporation data on civil legal needs and the Obermeyer et al. algorithmic audit [1,17]. Full derivation logic for all 11 parameters is documented in the public repository.

### Mathematical framework

Under the multiplicative barrier model, the probability of successful cascade completion is P(success) = ∏_i=1_ ^11^ p_i_. Writing the cascade as a product does not assume statistical independence: by the chain rule, P(S_1_,…,S_n_) = P(S_1_)∏_i≥2_ P(S_i_ | S_1_,…,S_i−1_), so the p_i_ are properly understood as stage-specific conditional pass probabilities. The effect of removing barrier j is Δ_j_ = P(success | p_j_ = 1) − P(success).

#### Proposition 1 (Single-barrier improvement bound)

Removing barrier j yields Δ_j_ = P · (1/p_j_ − 1). Thus absolute improvement is linear in P, the baseline success. When P ≈ 1.8 × 10^−5^, every Δ_j_ is necessarily small regardless of which barrier is targeted. This is an algebraic property of series systems, not a simulation finding.

#### Proposition 2 (Higher-order interaction under probability-scale multiplicativity)

Define layer aggregate probabilities L_k_ = ∏_i k_ p_i_ for layers k = 1, 2, 3, so that P = L_1_ · L_2_ · L_3_. On the probability scale, factorial contrasts of P generate nonzero higher-order (including three-way) interaction terms, whereas on the log scale log P = ∑ log p_i_ is additive and interactions vanish. The *existence* of higher-order interaction is thus a structural feature of probability-scale multiplicativity; its *magnitude* (e.g., the 87.6% three-way share) is parameterization- and topology-dependent and is not a universal property [8].

### Shapley value attribution

To fairly attribute system effect to individual barriers, we employed Shapley value decomposition from cooperative game theory [18], assigning each barrier its average marginal contribution across all possible removal orderings.

### Sensitivity analysis

We conducted comprehensive sensitivity analysis including one-at-a-time sensitivity (±10%), Sobol global sensitivity indices [19] estimated by Monte Carlo A/B (Saltelli/Jansen) sampling (n = 1,024 base samples) and Morris screening [20]. All barrier probabilities were perturbed across ±33% multiplicative ranges and across empirically bounded intervals. Parameter-perturbation resampling (n = 1,000) characterized the stability of the principal findings within these ranges.

To assess whether the conclusions depend on the serial, single-pass structure and on the substitution of marginal for conditional probabilities, we conducted two alternative-topology analyses (seed 42, n = 100,000). First, we coupled the 11 Bernoulli pass-events through a Gaussian copula with latent equicorrelation ρ (the Gaussian copula correlation parameter, not the post-thresholding Bernoulli correlation) swept from 0 to 0.5 (and a within-layer block variant), preserving each marginal pass probability. Second, we allowed m independent opportunities to clear each stage, p_i_^(m)^ = 1 − (1 − p_i_)^m^, for m = 1, 2, 3. For each we recomputed the baseline success probability, the maximum single-barrier and single-layer effects, and the three-way interaction share.

### Software and reproducibility

All analyses were conducted in Python 3.10+ using NumPy, SciPy and Matplotlib; Sobol and Morris indices were computed with custom implementations cross-checked against SALib [21]. All stochastic analyses use a fixed seed (42); a one-command ‘mak’ pipeline reproduces every result, table and figure, and regression tests pin the deterministic headline values. Code and data are archived at Zenodo (DOI:10.5281/zenodo.22312977) and available at https://github.com/Nyx-Dynamics/algorithmic-bias-epidemiology-academic.

### Use of artificial intelligence

Large language models (Anthropic Claude and OpenAI ChatGPT) were used for literature search support and language refinement. No generative-AI–generated images or figures were used; all figures were produced programmatically using Matplotlib. JetBrains Junie was utilized for code correction, Zotero AI for reference management and the manuscript was prepared using Overleaf AI for LaTeX code correction. Specific model versions were not systematically recorded at the time of use; only the tools and their roles are stated. All analyses, interpretation and conclusions were conducted independently by the author.

## Ethics statement

This study used publicly available, aggregate federal datasets and published peer-reviewed statistics. No individual-level data were collected or analyzed and no human subjects were enrolled. Institutional review board approval was not required.

## Acknowledgments

The author thanks the open-source software communities whose tools made this analysis possible. This work received no external funding.

## Author contributions

A.C.D. conceived the study, developed the mathematical framework, conducted all analyses, interpreted results and wrote the manuscript.

## Supporting information

**S1 File.** Supplementary Tables S1–S7 and Supplementary Figures S1–S3: individual barrier removal effects; Shapley value attribution (seeded); Sobol sensitivity indices (seeded); a bounded-perturbation stability summary; signal-to-noise and coefficient-of-variation analysis; a full parameter-derivation audit trail (Table S6); and alternative-topology robustness (Table S7), with figures for Shapley attribution, layer-removal effects, and alternative-topology robustness.

## Data availability statement

All code and data are archived at Zenodo (DOI:10.5281/zenodo.22312977), the citable revision artifact, with the development repository at https://github.com/Nyx-Dynamics/algorithmic-bias-epidemiology-academic. Derived data are provided as CSV files under `data/processed`.

## Code availability

All analysis code is archived at Zenodo (DOI:10.5281/zenodo.22312977) and available at https://github.com/Nyx-Dynamics/algorithmic-bias-epidemiology-academic. A one-command pipeline reproduces every result, table and figure: `make all` (full pipeline) and `make verify` (claims-vs-code checks and determinism), with regression tests pinning the deterministic headline values. Key scripts: `barrier_visualization.py` (model, counterfactuals, figures), `sensitivity_analysis.py` (Sobol/Morris and parameter-perturbation analysis), `copula_robustness.py` (alternative topologies), `systemic_bias_sensitivity.py`, and `build_derivation_table.py`. Environment: Python 3.10+, NumPy, SciPy, Matplotlib. Fixed seed: 42.

