## Supplemental Text and Tables for "Structural limits of single-barrier reform in algorithmic recourse: a formal series-system model with implications for digital health"

A.C. Demidont

*Scope of claims.* All numerical values below (the 0.0018% baseline, the 0.0054% maximum single-barrier gain, the 87.6% three-way share, and the Shapley/Sobol/bootstrap/SNR outputs) are model-derived illustrations under a provisional cross-domain parameterization; they are consistent with, but not independent confirmation of, Propositions 1–2. All stochastic analyses are seeded (seed 42) and reproducible via the repository make pipeline.

**Supplementary Table 1: Individual barrier removal effects.**

| Barrier | Layer | Baseline P (%) | P with removal (%) | Δ (%) | 95% CI |
| --- | --- | --- | --- | --- | --- |
| Rapid Data Transmission | L1 | 0.0018 | 0.0060 | 0.0042 | (0.0038, 0.0046) |
| Multi-System Integration | L1 | 0.0018 | 0.0033 | 0.0015 | (0.0013, 0.0017) |
| Permanent Storage | L1 | 0.0018 | 0.0040 | 0.0022 | (0.0020, 0.0024) |
| Error Detection Difficulty | L2 | 0.0018 | 0.0051 | 0.0033 | (0.0030, 0.0036) |
| Correction Process Barriers | L2 | 0.0018 | 0.0051 | 0.0033 | (0.0030, 0.0036) |
| Incomplete Correction Propagation | L2 | 0.0018 | 0.0045 | 0.0027 | (0.0024, 0.0030) |
| Awareness Gap | L3 | 0.0018 | 0.0060 | 0.0042 | (0.0038, 0.0046) |
| Record Access Barriers | L3 | 0.0018 | 0.0033 | 0.0015 | (0.0013, 0.0017) |
| Legal Knowledge Gap | L3 | 0.0018 | 0.0072 | 0.0054 | (0.0050, 0.0060) |
| Legal Resource Barriers | L3 | 0.0018 | 0.0045 | 0.0027 | (0.0024, 0.0030) |
| Systemic Bias in Algorithms | L3 | 0.0018 | 0.0060 | 0.0042 | (0.0038, 0.0046) |

L1 = Data Integration; L2 = Data Accuracy; L3 = Institutional. All individual effects are <0.02%, consistent with Proposition 1. CIs from bootstrap resampling (n = 1,000).

**Supplementary Table 2: Shapley value attribution.**

| Barrier | Layer | Shapley value (%) | Rank |
| --- | --- | --- | --- |
| Legal Knowledge Gap | Institutional | 10.95 | 1 |
| Rapid Data Transmission | Data Integration | 10.72 | 2 |
| Systemic Bias in Algorithms | Institutional | 10.53 | 3 |
| Incomplete Correction Propagation | Data Accuracy | 10.10 | 4 |
| Awareness Gap | Institutional | 9.67 | 5 |
| Error Detection Difficulty | Data Accuracy | 9.53 | 6 |
| Correction Process Barriers | Data Accuracy | 9.21 | 7 |
| Legal Resource Barriers | Institutional | 8.50 | 8 |
| Permanent Storage | Data Integration | 7.79 | 9 |
| Record Access Barriers | Institutional | 6.79 | 10 |
| Multi-System Integration | Data Integration | 6.21 | 11 |

Shapley values account for interaction effects across all possible removal orderings, providing fair attribution even under multiplicative blocking.

**Supplementary Table 3: Sobol sensitivity indices.**

| Barrier | S1 | ST | Interaction (ST−S1) |
| --- | --- | --- | --- |
| Rapid Data Transmission | 0.063 | 0.101 | 0.038 |
| Multi-System Integration | 0.073 | 0.103 | 0.030 |
| Permanent Storage | 0.059 | 0.097 | 0.038 |
| Error Detection Difficulty | 0.069 | 0.098 | 0.029 |
| Correction Process Barriers | 0.088 | 0.103 | 0.015 |
| Incomplete Correction Propagation | 0.074 | 0.101 | 0.027 |
| Awareness Gap | 0.075 | 0.100 | 0.025 |
| Record Access Barriers | 0.075 | 0.108 | 0.033 |
| Legal Knowledge Gap | 0.084 | 0.103 | 0.019 |
| Legal Resource Barriers | 0.094 | 0.106 | 0.011 |
| Systemic Bias in Algorithms | 0.065 | 0.098 | 0.034 |

S1 = first-order (main) effects; ST = total effects including interactions. Saltelli sampling with n = 1,024 base samples (seeded). ST exceeded S1 for all barriers, but the gaps are small and, for several barriers, within Monte-Carlo error at this sample size; the characterization “substantially exceeds S1 for all barriers” is therefore not supported.

**Supplementary Table 4: Bootstrap robustness summary.**

| Finding | Threshold | % resamples supporting |
| --- | --- | --- |
| Three-way factorial share (>70% of achievable improvement) | >70% | 100.0% |
| Maximum individual barrier effect | <1% | 100.0% |
| Barriers needed for 90% success | ≥10 of 11 | 100.0% |

Parameter-perturbation resampling with n = 1,000 resamples and ±10% perturbation. Computed with the identical baseline factorial decomposition (subtracting main and pairwise terms), the three-way factorial share under perturbation had mean 87.7% (95% CI 84.8–90.3), centered on the canonical 87.6% — i.e., the interaction structure is stable under ±10% perturbation.

**Supplementary Table 5: Signal-to-noise ratio analysis.**

| Noise level (%) | SNR (dB) | Quality | CV (%) |
| --- | --- | --- | --- |
| 1 | 14.8 | Good (>10 dB) | 3.3 |
| 3 | 11.7 | Good (>10 dB) | 6.8 |
| 5 | 7.8 | Marginal (3–10 dB) | 16.6 |
| 10 | 4.6 | Marginal (3–10 dB) | 35.1 |
| 15 | 2.8 | Marginal (3–10 dB) | 53.0 |
| 20 | 1.4 | Poor (<3 dB) | 73.0 |
| 25 | −0.4 | Poor (<0 dB) | 109.8 |
| 30 | −1.1 | Poor (<0 dB) | 128.7 |

SNR remains positive (>0 dB) up to approximately 20% parameter noise (seeded); the coefficient of variation rises to 109.8% at 25% and 128.7% at 30% noise, so robustness is bounded and is not claimed under arbitrary uncertainty. CV = coefficient of variation of output success probability.

**Supplementary Table 6: Parameter derivation audit trail (all 11 barriers).**

For each barrier: source domain, exact source statistic, resulting pass probability, plausibility range, and transportability limitation. This makes each parameter auditable from the manuscript package alone; the full derivation logic is mirrored in the repository (data/parameter_sources/parameter_derivation.md). Each statistic is drawn from the cited primary source and year.

| Barrier (layer) | Source domain | Source statistic [source] | p | Range | Transportability limitation |
| --- | --- | --- | --- | --- | --- |
| Rapid Data Transmission (L1) | Credit reporting | Furnishers transmit adverse data to CRAs within 1–2 days [CFPB 2022] | 0.30 | 0.20–0.40 | Credit-furnishing timing; not measured for clinical/EHR data flows |
| Multi-System Integration (L1) | Credit reporting | 20% had error on ≥1 of 3 reports [FTC 2013] | 0.55 | 0.40–0.65 | Cross-bureau error; not clinical multi-system integration |
| Permanent Storage (L1) | Credit reporting | 7-yr retention; <20% removed in 4 yr [CFPB 2022; FCRA §605] | 0.45 | 0.35–0.55 | FCRA retention regime; not healthcare record retention |
| Error Detection Difficulty (L2) | Credit reporting | 26% identified material errors [FTC 2013] | 0.35 | 0.25–0.45 | Consumer credit review; not clinical error detection |
| Correction Process Barriers (L2) | Credit reporting | 37% of disputes fully resolved [FTC 2013; CFPB 2022] | 0.35 | 0.25–0.45 | Credit dispute process; not clinical data correction |
| Incomplete Correction Propagation (L2) | Credit reporting | Corrections do not auto-propagate across CRAs [CFPB 2022; FTC 2015] | 0.40 | 0.30–0.50 | Inter-CRA propagation; not clinical systems |
| Awareness Gap (L3) | Civil legal | Help sought for ~1 in 4 civil legal problems [LSC 2022] | 0.30 | 0.20–0.40 | Civil legal need; not algorithmic-decision awareness |
| Record Access Barriers (L3) | Credit reporting | Statutory free-report access with practical barriers [FCRA §612; CFPB 2022] | 0.55 | 0.45–0.65 | FCRA access; not clinical record access |
| Legal Knowledge Gap (L3) | Civil legal | 39% believe they can use the legal system [LSC 2022] | 0.25 | 0.15–0.35 | General legal confidence; not algorithm-specific rights |
| Legal Resource Barriers (L3) | Civil legal | 46% cite cost; ~50% of applicants turned away [LSC 2022] | 0.40 | 0.30–0.50 | Civil legal-aid capacity; not algorithm-specific representation |
| Systemic Bias in Algorithms (L3) | Healthcare audit | Cost-for-need proxy bias; correcting it raises Black enrollment 17.7%→46.5% [Obermeyer 2019] | 0.30 | 0.20–0.40 | Single commercial tool / one health system; NOT a population-wide bias rate (a prior “62%” figure was erroneous and is corrected) |

**Supplementary Table 7: Robustness to alternative topologies (seed 42, n = 100,000).**

| Scenario | Baseline P | 3-way share | max single-barrier | max single-layer |
| --- | --- | --- | --- | --- |
| Independence (ρ = 0, analytic reference) | 0.0018% | 87.6% | 0.0054% | 0.36% |
| Copula ρ = 0.1 | 0.049% | 84.5% | 0.071% | 0.97% |
| Copula ρ = 0.2 | 0.288% | 81.9% | 0.185% | 1.85% |
| Copula ρ = 0.3 | 0.835% | 79.9% | 0.376% | 2.74% |
| Copula ρ = 0.4 | 1.778% | 78.5% | 0.606% | 3.71% |
| Copula ρ = 0.5 | 3.200% | 77.4% | 0.821% | 4.50% |
| Block copula ρ = 0.5 | 0.151% | 66.0% | 0.142% | — |
| Repeated attempts m = 2 | 0.352% | 53.3% | 0.452% | 5.70% |
| Repeated attempts m = 3 | 3.651% | 25.2% | 2.665% | 16.88% |

Under correlation the comparative conclusion holds (single-layer ≪ coordinated) while the baseline value is correlation-sensitive; under repeated attempts interaction dominance attenuates, delimiting the model’s domain of validity to approximately single-pass serial recourse. ρ denotes the latent Gaussian-copula correlation parameter, not the post-thresholding Bernoulli correlation. The ρ = 0 row reports exact analytic values; the Monte-Carlo estimate at ρ = 0 is noise-limited (~2 successes per 10^5 draws) and appears in the repository CSV, so the low-ρ rows (ρ ≤ 0.15) carry visible Monte-Carlo error.

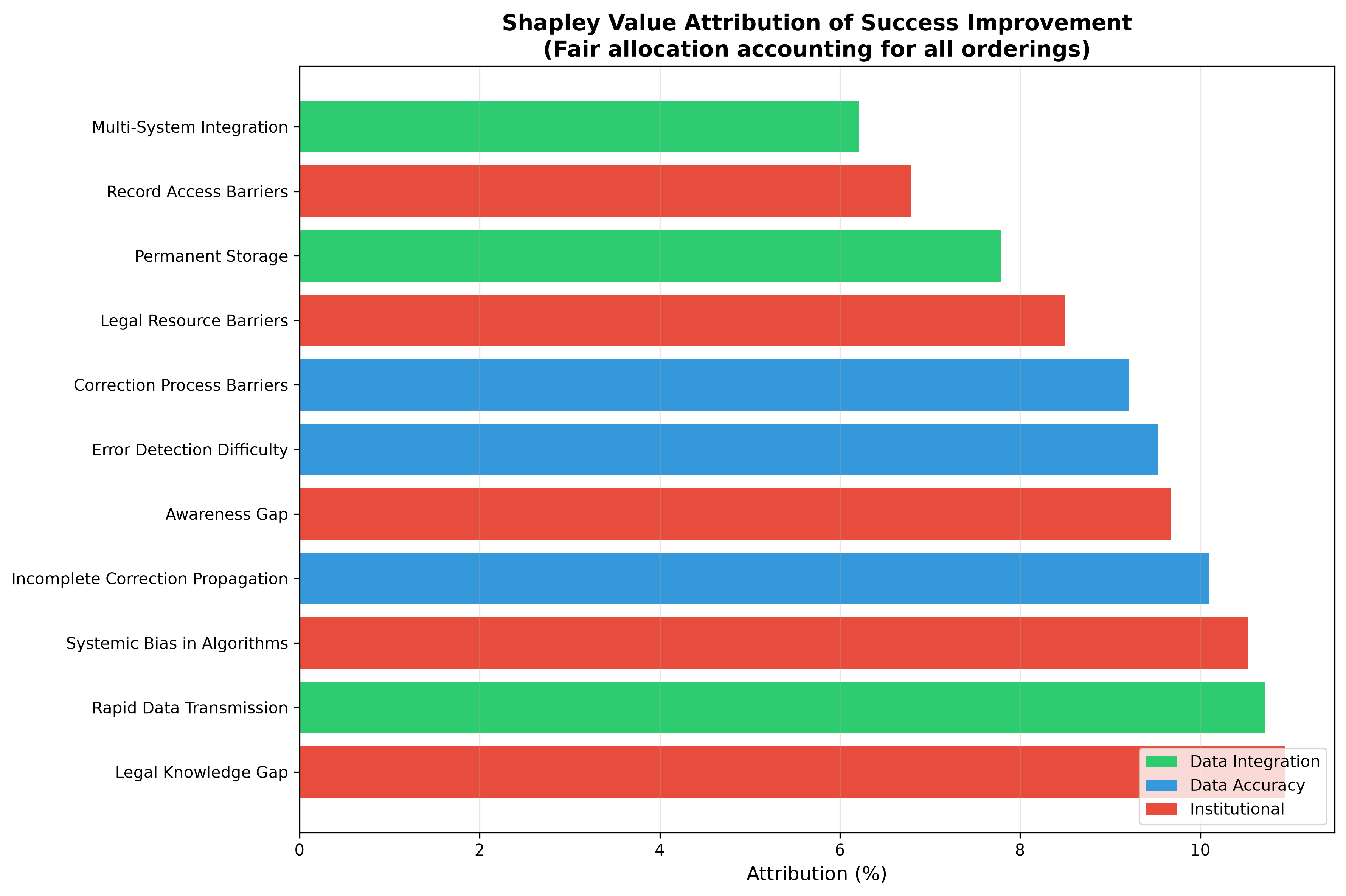

**Supplementary Figure 1: Shapley value attribution of barrier contributions.** Shapley value decomposition assigning relative contribution of each barrier to overall system success while accounting for all possible barrier removal orderings. Barriers are coloured by layer: green = Data Integration, blue = Data Accuracy, red = Institutional. Values represent fair attribution of total achievable improvement, with the five highest-contributing barriers spanning all three layers, consistent with the cross-layer nature of the modeled dynamics.

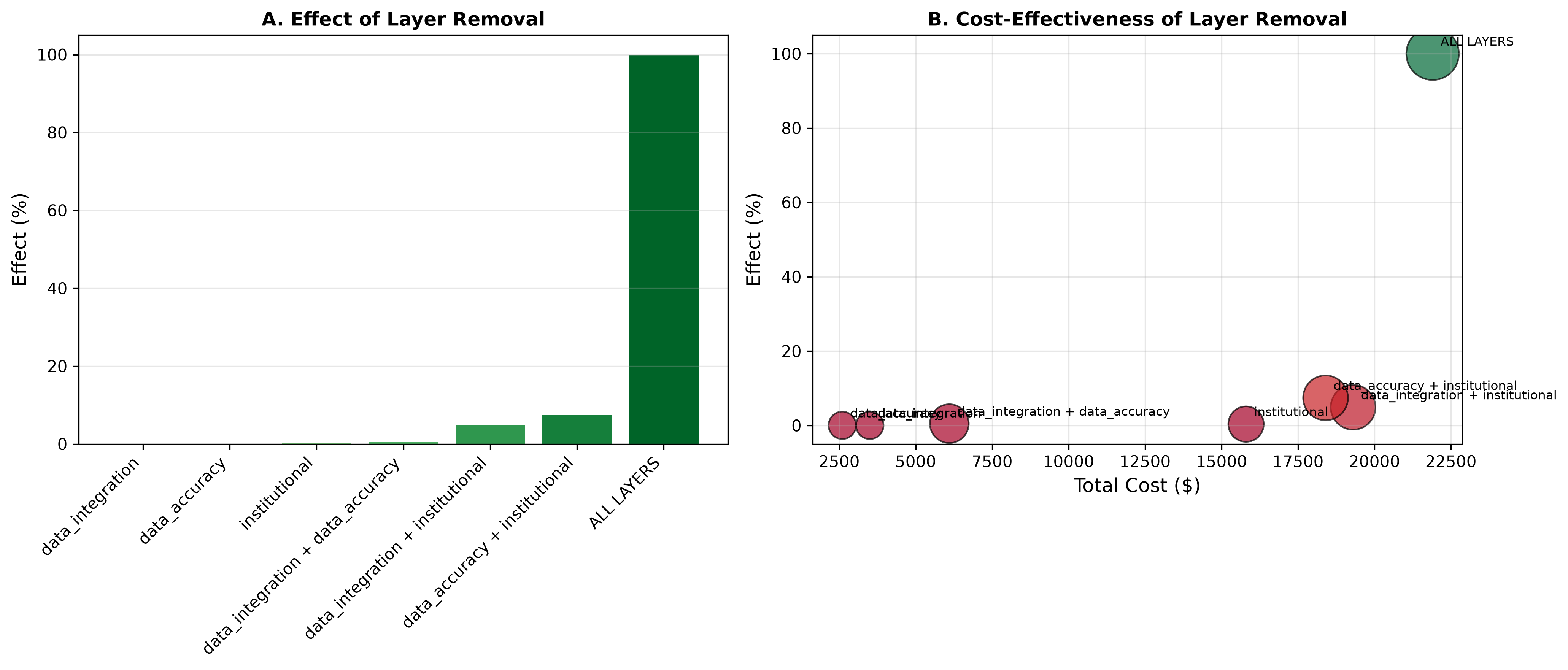

**Supplementary Figure 2: Effect of layer removal on system success probability.** Only complete removal across all three layers yields substantial improvement (100%). Single-layer removal produces negligible effects; two-layer combinations yield at most 7.4% (Data Accuracy + Institutional), consistent with the model’s three-way interaction structure.

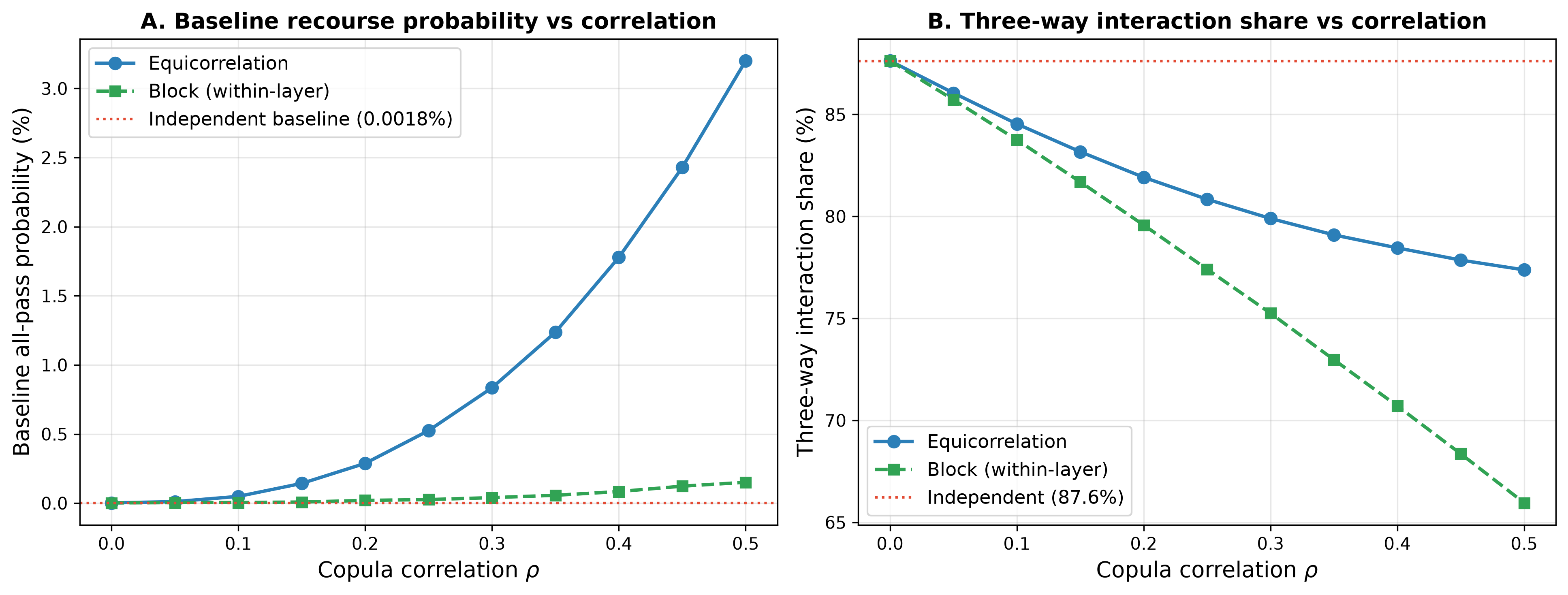

**Supplementary Figure 3: Robustness to alternative topologies.** Baseline success probability P (left) and three-way interaction share (right) as a function of latent Gaussian-copula correlation ρ, for the equicorrelation and within-layer block variants. The comparative conclusion (single-layer ≪ coordinated) is robust to correlation while the baseline value is correlation-sensitive. The repeated-attempt results (m = 2, 3), which attenuate interaction dominance, are reported in Supplementary Table 7.
