## Supplementary figures and images for "Structural limits of single-barrier reform in algorithmic recourse: a formal series-system model with implications for digital health"

### figure S1

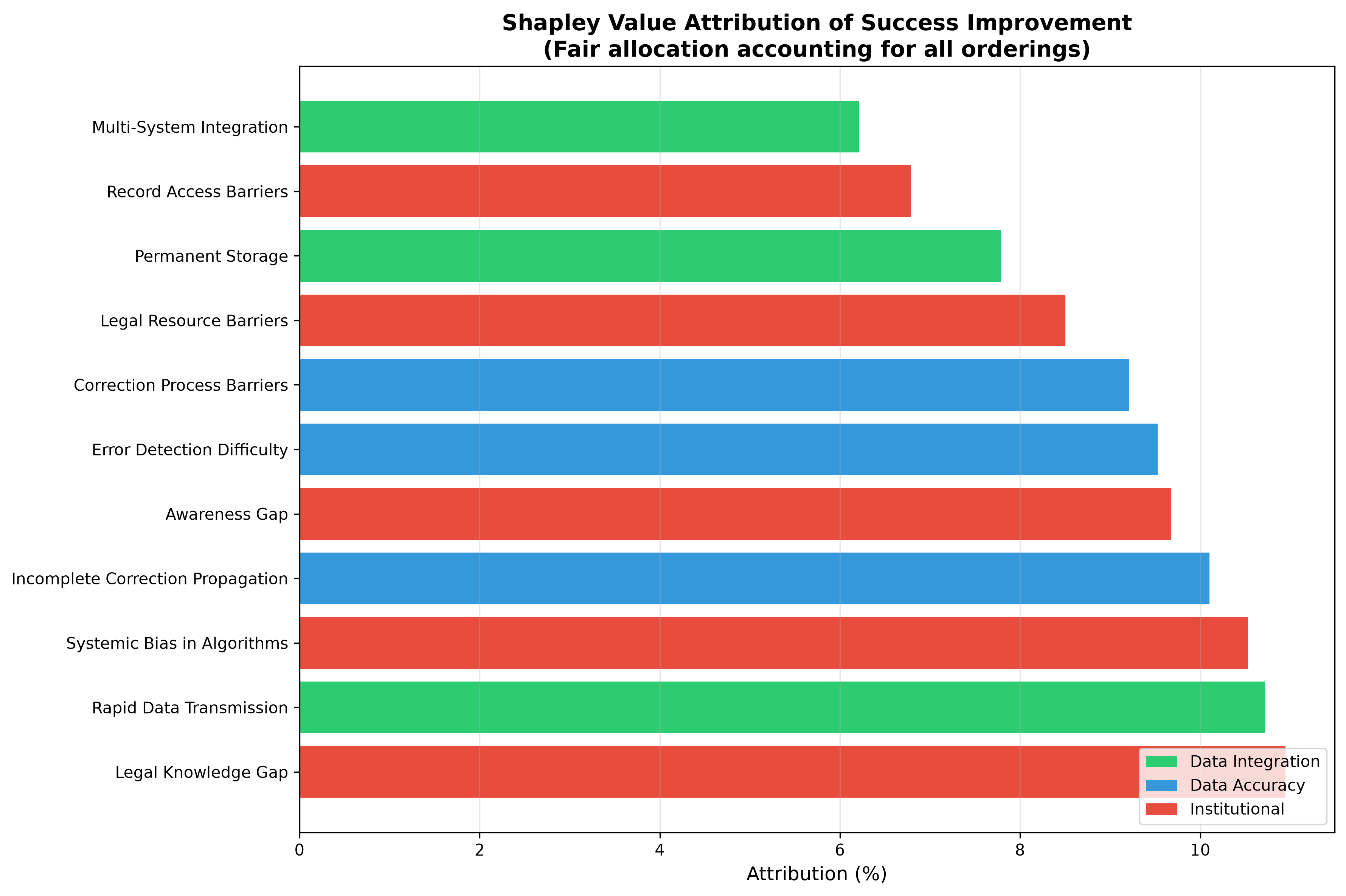

### figure S2

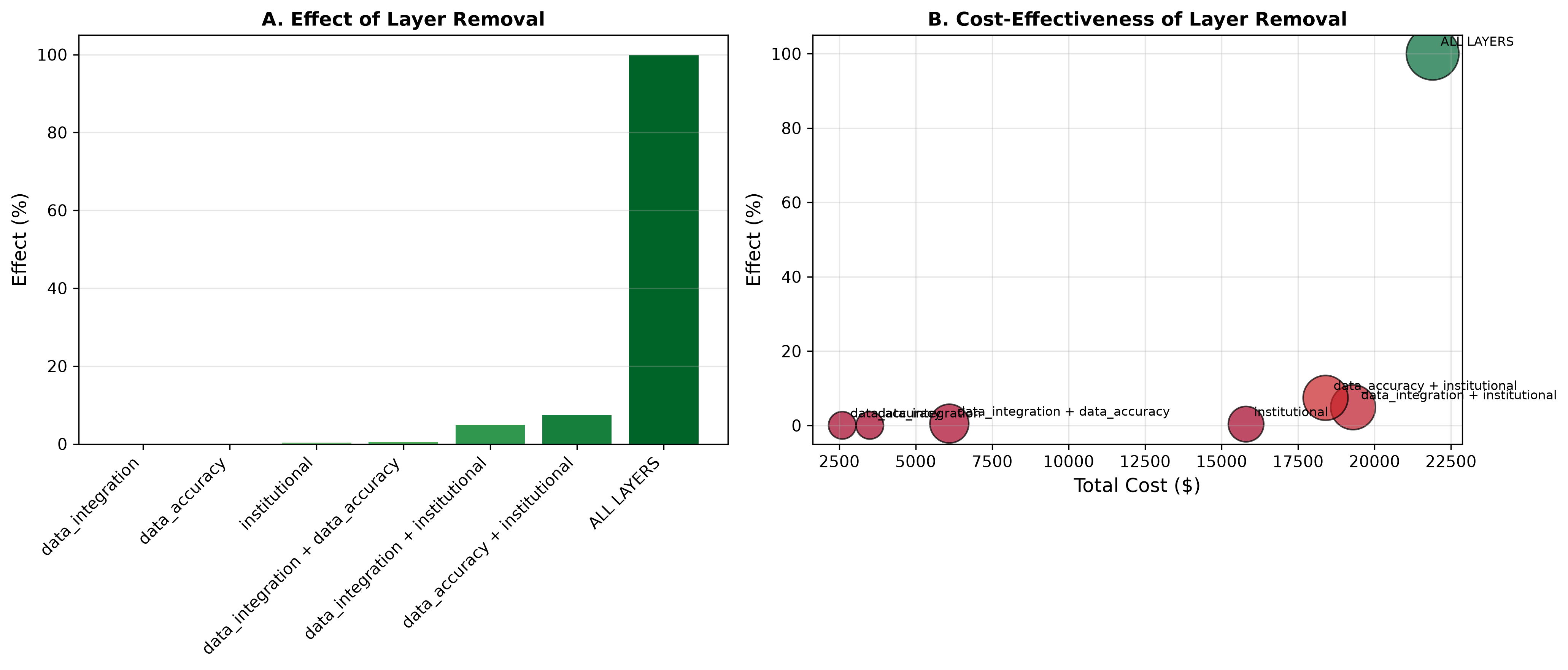

### figure S3

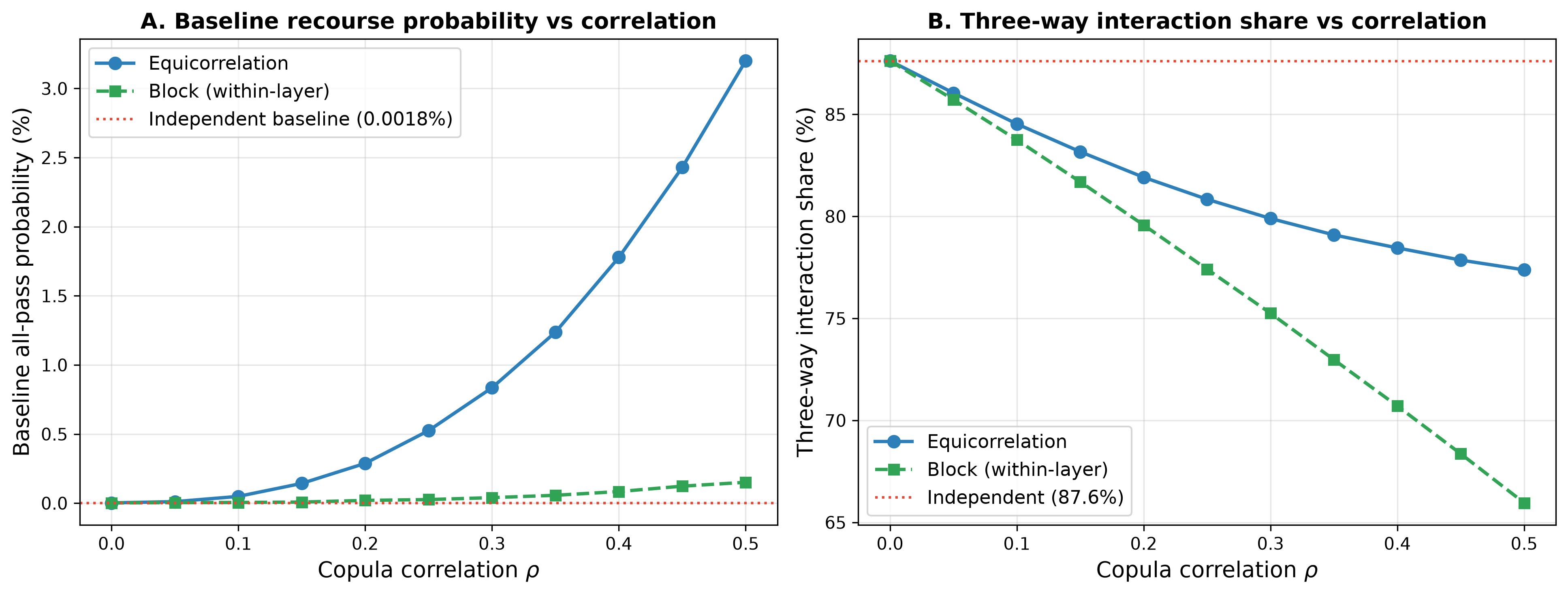
